# Low levels of systemic inflammation in asymptomatic TB detected during community-based screening in rural South Africa

**DOI:** 10.64898/2025.12.10.25341906

**Authors:** Lerato Mtshali, Stephen Olivier, Jana Fehr, Thando Zulu, Dickman Gareta, Kathy Baisley, Willem Hanekom, Thumbi Ndung’u, Alasdair Leslie, Emily B. Wong

## Abstract

Asymptomatic TB represents half of prevalent TB yet frequently remains undetected. We assessed the performance of plasma IL-6 and CRP as diagnostic biomarkers for asymptomatic TB identified by community-based screening. Both proteins were elevated in asymptomatic TB compared to community controls, but neither performed well as a diagnostic biomarker.

## INTRODUCTION

Tuberculosis (TB) remains the leading cause of infectious death worldwide [1]. In October 2024, the World Health Organization (WHO) defined bacteriologically-confirmed asymptomatic TB as disease featuring detectable Mycobacterium tuberculosis (M.tb) in a person who presents without symptoms at screening. [1]. Evidence from national TB prevalence surveys suggests that people with asymptomatic TB account for over half of all bacteriologically-confirmed prevalent TB [2, 3]. Identifying and treating these “missing” TB cases through community-based screening may be crucial to halting transmission and decreasing TB morbidity and mortality.

Elevated plasma concentrations of inflammatory proteins have shown promise as biomarkers for diagnosing TB and monitoring treatment response [4, 5]. WHO recommends the use of elevated CRP as a screening biomarker for people living with HIV (PLWH) [6]. However, this guidance is based on findings from symptomatic TB detected in clinical settings and its utility in community-based screening for asymptomatic TB remains unclear.

## MATERIALS AND METHODS

This nested case-control study used stored plasma samples from participants in the Vukuzazi, a community-based multi-disease survey[7] and the Regional Prospective Observational Research for Tuberculosis - South Africa (RePORT-SA) study (detailed in Supplementary Methods). Individuals from Vukuzazi meeting the following definitions for bacteriologically-confirmed TB were included: community-detected asymptomatic TB (negative WHO 4 symptom screen (W4SS) and M.tb sputum-positive (by GeneXpert Ultra and/or liquid mycobacterial culture); community-detected symptomatic TB (positive W4SS and M.tb sputum-positive). For comparison, people with clinic-diagnosed symptomatic TB (positive W4SS and M.tb sputum-positive) and community controls (negative W4SS, normal chest X-ray and sputum-negative) were randomly selected from RePORT-SA and Vukuzazi respectively and frequency-matched for age, sex and HIV-status to the asymptomatic TB group.

Plasma CRP and IL-6 concentrations (measured by fluorescent bead immunoassay) were compared between participant groups and stratified by HIV-status (Median [Interquartile range], Mann-Whitney U-test). Measure of HIV (ART status, viral load and CD4 counts) and TB (bacterial and radiological burden) severity were also compared between groups. The diagnostic performance of each protein was assessed by receiver operator (ROC) curves, noting the sensitivity and specificity of CRP at a cut-off of 5mg/L (WHO recommended threshold for PLWH). Correlations between CRP, IL-6, bacterial burden (defined as 42 – number of days to positive liquid mycobacterial culture), and radiological severity (CAD4TBv7) were assessed using Spearman correlation.

Sensitivity analyses addressed uncertainties in GeneXpert Ultra “trace” results by using alternative definitions of community-detected bacteriologically-confirmed TB: (1) excluding people with GeneXpert Ultra “trace”-positive and culture-negative sputum and, (2) including only people with positive sputum by liquid mycobacterial culture (regardless of GeneXpert Ultra status). Statistical analyses were performed in GraphPad Prism 10.2.3 and STATA 18.

## RESULTS

The analysis included 302 participants: community-detected asymptomatic TB (N=142), community-detected symptomatic TB (N=30), clinic-diagnosed symptomatic TB (N=30), and community controls (N=100). The median age of asymptomatic TB participants was 42 years [31-62], 79 (56%) were female and 58 (41%) were PLWH. Other groups had similar demographic factors (Supplementary Table 1). Among people with asymptomatic TB who were PLWH, 46 (79%) were on ART, 39 (67%) had undetectable viral load and the median CD4 count of 566 [IQR 372-720]. Although, percentage on ART was similar (62%) among people with clinic-diagnosed TB, the median CD4 count was lower (382 [182-717]). The median bacterial burden in community-detected asymptomatic TB participants was 24 days [0-28] and the median CAD4TBv7 score was 57 [25-82]. Measures of disease severity were similar in community-detected symptomatic TB and more severe in clinic-detected symptomatic TB (Supplementary Table 1). For the first and second alternative definitions of bacteriologically-confirmed TB, 25 and 22 symptomatic and 108 and 91 asymptomatic participants met the definitions respectively (Supplementary Figure 1).

IL-6 and CRP concentrations were higher in asymptomatic TB, yellow, (Figure 1A) than in community controls, blue, (IL-6: 0.79pg/ml [0.04-1.91] vs 0.12pg/ml [0.04-1.06], p=0.0111); CRP: 5mg/L [1-13.5] vs 2mg/L [1-5], p= 0.0233), but lower than in clinic-diagnosed TB (maroon, IL-6: 11.31pg/ml [4.77-17.21], p<0.0001 and CRP: 88mg/L, [42.75-153.5], p<0.0001). Cytokine concentrations in symptomatic community-detected TB (orange, IL-6: 0.79pg/ml [0.078-3.5] and CRP: 7mg/L [2.75-26]) were similar to asymptomatic TB. Among people with asymptomatic TB, IL-6 and CRP levels did not differ significantly by HIV status (Supplementary Figure 2, IL6: 0.79mg/L [0.07-1.88] vs 0.66mg/L [0.04-2.07]; CRP 3mg/L [1-10] vs 8mg/L [1-17]). These findings were robust to the alternative definitions of bacteriologically-confirmed TB (Supplementary Figures 3&4).

IL-6 and CRP effectively distinguished clinic-diagnosed (maroon) TB from community controls (IL-6: AUC 0.87 (95% CI 0.7789-0.9650); and CRP: 0.97 (95% CI 0.9447-1.00)), but poorly distinguished asymptomatic TB (yellow) from community controls (IL-6: AUC 0.62 (95% CI 0.5541-0.6951) and CRP: 0.62 (95% CI 0.5456-0.6873)) (Figure 1B). At a CRP threshold of 5mg/L, the sensitivity and specificity for detecting asymptomatic TB were 51.1% and 69.4% respectively, which was similar to 56.7% sensitivity and 69.4% specificity for community-detected symptomatic TB, both much lower than the 96.2% sensitivity with comparable specificity (69.4%) for clinic-diagnosed TB (Supplementary Table 2). CRP performed better in PLWH than in people without HIV in both asymptomatic (AUC 0.66 (95% CI 0.5470-0.7629) vs. 0.60 (95% CI 0.5023-0.6922), p <0.0001) and symptomatic community-detected TB (AUC 0.81 (95% CI 0.6726-0.9411 vs 0.57 (95% CI 0.3875-0.7513), p <0.0001, Supplementary Figure 5). Sensitivity analyses indicated slight improvements in the AUC for asymptomatic TB for alternative definition 1 (IL-6: 0.64 (95% CI 0.5625-0.7128); CRP: 0.66 (95% CI 0.5819-0.7327)) and alternative definition 2 (IL-6 0.65 (95% CI 0.5684 to 0.7263); CRP: 0.67 (95% CI 0.5860-0.7449), Supplementary Figures 6A&B, 7A&B and Supplementary Table 2.

**Figure 1.**
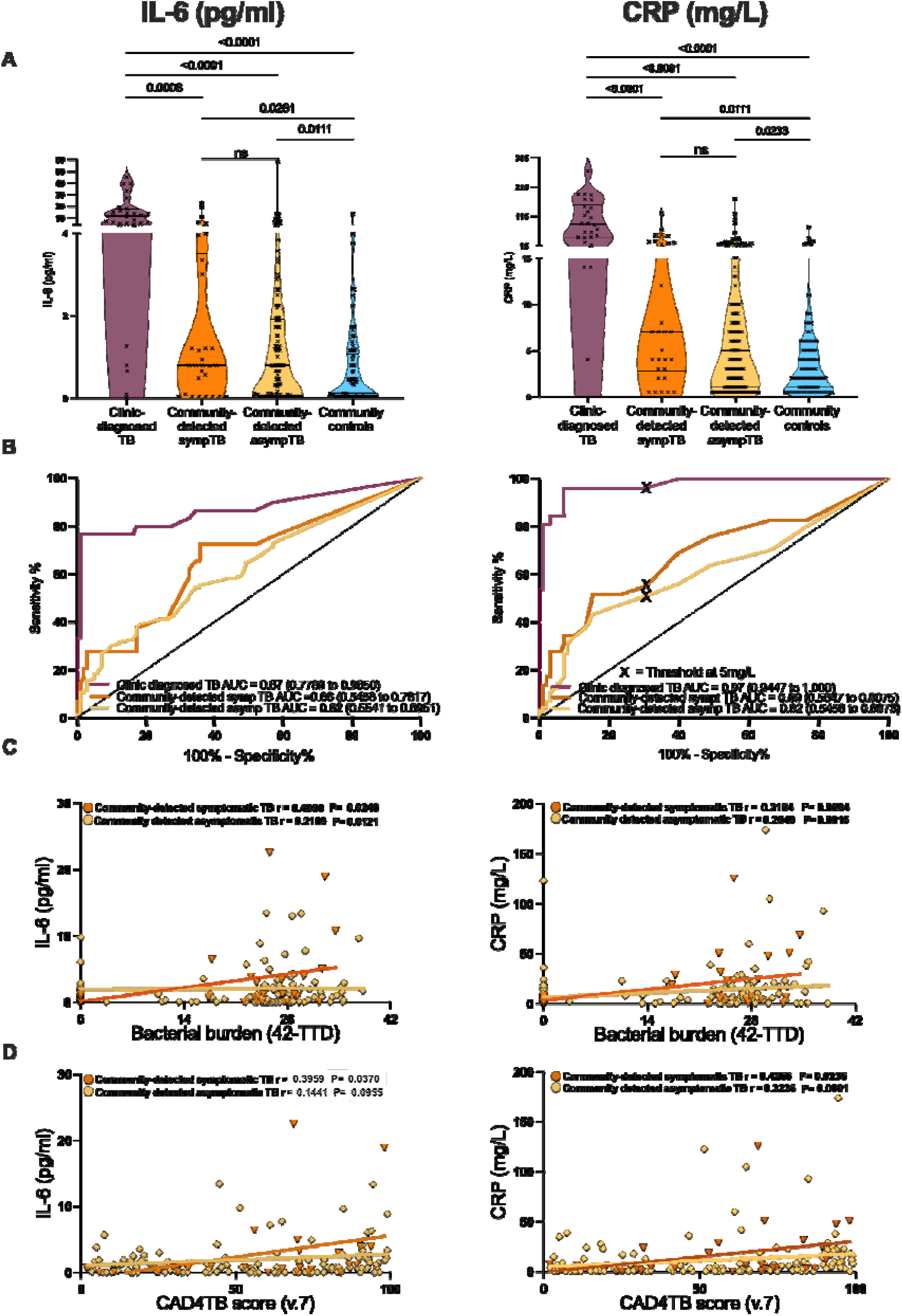
(A) Differences in plasma concentrations of IL-6 and CRP measured by fluorescent bead assay in clinic-diagnosed TB (maroon), community-detected symptomatic TB (orange), community-detected asymptomatic TB (yellow) and community controls (blue). Data show median values and IQR, P-values was obtained using Mann-Whitney U test for a nonparametric dataset. (B) Receiver operator curve assessing the performance of IL-6 and CRP as biomarkers for distinguishing clinic-diagnosed TB (maroon), community-detected symptomatic TB (orange) and community-detected asymptomatic TB (yellow) from community controls. Data show area under the curve (AUC) and the 95% confidence interval. ‘X’ represents the sensitivity and specificity at CRP threshold 5mg/L, chosen based on WHO’s recommendation for required triage test. (C-D) Correlation analysis between people with asymptomatic (yellow) and symptomatic community-detected TB (orange), and plasma IL-6 and CRP concentrations with (C) bacterial burden (42 minus Time to Detection (days) (D) extent of lung involvement measured by CAD4TBv7 scores, determined from digital chest X-rays. Statistically significant differences are reported.

In asymptomatic TB (yellow), both inflammatory proteins weakly but significantly correlated with bacterial burden (IL-6: r=0.2109, p=0.0121; CRP: r=0.2646, p=0.0015, Figure 1C). This correlation was stronger in clinic-diagnosed TB (IL-6: r=-0.3627, p=0.0489; CRP: r=0.5385, p=0.0045) and symptomatic community-detected TB (orange) for IL-6 (r=0.4090, p=0.0248) but not for CRP (r=0.3184, p=0.0864, Supplementary Figure 8A&B). In community-detected asymptomatic TB, CRP correlated with radiological burden (r=0.3263, p=0.0001), while IL-6 did not (r=0.1441, p=0.0955). These results were consistent across sensitivity analyses (Supplementary 9&10).

## DISCUSSION

In this study, plasma concentrations of IL-6 and CRP, were higher in people with asymptomatic community-detected TB than community controls, but much lower than in people with clinic-diagnosed TB. CRP is an established biomarker of inflammation and a recommended tool for TB screening among PLWH [4, 8]. In this study, CRP poorly distinguished asymptomatic TB from community controls, falling far short of the WHO target for a triage test with 90% sensitivity and 70% specificity [9]. These results confirm the findings of previous studies that show CRP is a poor biomarker for community-detected asymptomatic TB [10-12]. Although CRP demonstrated somewhat superior performance in PLWH, it still had poor sensitivity (61.4%) and specificity (64.4%) for detecting asymptomatic TB at the 5mg/L threshold. This study also found that IL-6, another inflammatory cytokine that has been shown at high concentrations to be associated with TB disease severity and increased lung pathology, is somewhat elevated in community-detected asymptomatic TB but does not have sufficient discriminatory performance to serve as a screening tool to distinguish asymptomatic TB from healthy controls.

An important new finding from our work is that levels of inflammation between symptomatic and asymptomatic TB detected by community-based screening did not differ from each other. Stratifying by symptom presentation only slightly increased CRP sensitivities from 51.1% to 56.7%. This finding contributes to a discussion in the field regarding the ‘frailty’ of symptom reporting for TB screening, and suggests that TB detected during screening, regardless of symptom status at the time of screening is biologically more ‘mild’ than symptomatic TB that is detected as a result of passive case finding.

The people with asymptomatic TB in this study demonstrated a wide spectrum of systemic inflammation that included substantial numbers of individuals with undetectable levels of CRP and IL-6 as well as individuals with levels as high as those observed in clinic-diagnosed TB. This heterogeneity is only partially explained by weak correlations with measures of bacteriologic or radiologic severity, suggesting that further work must be done to understand the biological heterogeneity of early TB disease.

While this study provides important insights, there were several limitations. First, symptoms were self-reported and not extensively characterized. Secondly, universal sputum collection was not performed and for those triaged for sputum collection only one sample was taken; both of these factors may have resulted in individuals with asymptomatic TB and normal chest x-rays or very pauci-bacillary disease being missed. Thirdly, the study captured data at a single time-point and in an HIV-endemic population in rural KwaZulu-Natal limiting generalizability. In addition, in the primary definition of bacteriologically-confirmed TB, which included GeneXpert trace, may have been overly permissive including some individuals with false-positive results who did not really have bacteriologically-confirmed active TB; nonetheless the key findings were confirmed through more bacteriologically-stringent alternative definitions.

Taken together these findings suggest that community-detected TB, regardless of symptoms, may represent an earlier or milder phenotype of TB, defined by low yet heterogeneous levels of inflammation. These findings indicate that diagnostic biomarkers identified for their performance in advanced TB disease have limited utility to identify asymptomatic TB in the community. These results highlight the need for the identification of more effective and inexpensive diagnostic tests to identify people in this group.

## Data Availability

All data produced in the present work are contained in the manuscript

## Author contributions

Mtshali L.N. and Wong E.B. conceived and designed the study, with Mtshali L.N., Wong E.B., and Baisley K. developing the methodology and study protocol. Mtshali L.N. led the laboratory work, coordinated data collection, performed data cleaning, conducted statistical analyses, and interpreted the results, integrating input from all co-authors. Zulu T., and Gareta D., respectively supported sample preparation and data collation. Olivier S. supported data collection and assisted with statistical analyses. Mtshali L.N. drafted the manuscript and Fehr J., Leslie A., and Hanekom W. provided critical reviews and intellectual input in later drafts. Ndung’u T. co-supervised the study, and Wong E.B. provided overall supervision, funding, and project oversight. All authors approved the final manuscript.

