## Supplemental Material for "Low levels of systemic inflammation in asymptomatic TB detected during community-based screening in rural South Africa"

SUPPLEMENTARY MATERIALS

### SUPPLEMENTARY METHODS

#### Vukuzazi study participants

Vukuzazi is a comprehensive, community-based, cross-sectional screening survey carried out in the uMkhanyakude District of Kwa-Zulu Natal between May 25, 2018, and March 13, 2020. A total of 36,097 residents aged 15 years or older within the survey area were deemed eligible for participation and were invited to mobile health camps. The purpose of these camps was to conduct a health survey aimed at assessing the burden and intersection of HIV, tuberculosis, hypertension, and diabetes. Additionally, samples were collected for biobanking purposes.

TB screening was performed via a symptom presentation assessment as well through digital chest X-radiography for all non-pregnant participants. Those who were positive on the WHO 4 symptom screen (W4SS) or had abnormal lung pathology, as determined by computer-assisted image analysis (CAD4TBv5 $\geq$ 25) or an expert radiologists, were required to provide sputum samples. These samples were then tested for M.tb using the Xpert® MTB/RIF Ultra test (Cepheid, Sunnyvale, USA) and/or liquid mycobacterial culture (BACTEC™ MGIT™ 960 System, Becton Dickinson, Berkshire, UK). Whole blood samples obtained via venipuncture were collected for plasma separation, blood smears and HIV testing using the Genscreen Ultra HIV Ag-Ab enzyme immunoassay (Bio-Rad).

#### RePORT-SA study participants

The Regional Prospective Observational Research for Tuberculosis (RePORT) - South Africa study was conducted in Durban, South Africa, from December 2016 to December 2019. Participants were identified through passive case finding, were 18 years or older, and provided written informed consent. Individuals presenting with TB symptoms (positive W4SS) were referred for diagnostic testing by their attending clinicians. Pariticipants who tested positive for M.tb by GeneXpert and sputum culture (liquid and/or solid media) were enrolled in the study.

#### Measures of HIV and TB severity

Available data from both studies included: age, sex, HIV status, time to M.tb culture positivity, which were used to calculate bacterial burden (42 - time to detection, in days), and severity of chest x-ray abnormality (CAD4TBv7, Delft, Netherlands).

#### Measurement of IL-6 and CRP

Using the Luminex assay, we measured the levels of Interluekin-6, S100A9 and IL-1beta and C-Reactive Protein (R&D systems, Canada) according to the manufacturers instructions. Briefly, mangnetic beads coated with protein-specific antibodies were incubated with prediluted plasma samples at (1:2) for IL-6, S100A9 and IL-1beta and 1:200 for CRP in a 96-well plate. Proteins bound to beads are then incubated with detection antibodies (Biotin) with a and a streptavidin-phycoerythrin conjugate that can be read using the Bio-Plex 200 systems platform (Bio-Rad, USA). Bio-Plex Manager Software V.621 was then used for bead acquisition and analysis.

#### Sample size calculation

All available community-detected TB samples were utilized; the sample size for the comparison groups was calculated to have 80% power to demonstrate difference between symptomatic and asymptomatic TB based on the assumption that IL-6 levels in asymptomatic TB would be similar to people with symptomatic TB who had received effective treatment for two months (effect size 1.9) [13].

#### SUPPLEMENTARY TABLES

Table 1: Study participants characteristics of clinic-diagnosed TB, community-detected TB and community controls.

| Characteristics | N | Clinic-diagnosed TB N = 30[1] | Community-detected symptomatic TB N = 30[1] | Community-detected asymptomatic TB N = 142[1] | Community controls N = 100[1] | p-value[2] |
|---|---|---|---|---|---|---|
| Demographic data | | | | | | |
| Female, N (%) | 302 | 11 (37%) | 13 (43) | 79 (56) | 53 (53) | 0.2 |
| Age, years, median (IQR) | 302 | 36 [25; 46] | 54 [42; 66] | 42 [31; 62] | 49 [28; 62] | - |
| HIV positive, N (%) | 302 | 13 (43%) | 16 (53%) | 58 (41%) | 44 (44%) | 0.6759 |
| On ART , N (%) | 125 | 8(62%) | 12 (75%) | 46 (79%) | 34(77%) | <0.001 |
| CD4 count (IQR) | 132 | 382 [182; 717] | 592 [350; 919] | 566 [372; 720] | 533 [412; 717] | 0.2 |
| Virally Suppressed[a] , N (%) | 132 | 7 (54%) | 10 (63%) | 39 (67%) | 32 (73%) | 0.7 |
| Bacterial burden | | | | | | |
| Sputum GeneXpert Positive, N (%) | 172 | | 20 (67%) | 118 (83%) | - | 0.040 |
| Trace | 138 | - | 8 (40%) | 52 (44%) | - | 0.7 |
| Unknown | | - | 10 | 24 | - | |
| Time to detection (TTD) of M.tb on MGIT culture (days), median (IQR) | 202 | 8 [7; 11] | 18 [14; 42] | 18 [14; 42] | - | <0.0001 |
| Bacterial Burden (42 days-TTD) | 202 | 34 [31; 35] | 24 [0; 28] | 24 [0; 28] | | <0.0001 |
| Radiological burden | | | | | | |
| CAD4TB scores v7, median (IQR) | 286 | 74 [58; 90] | 70 [31; 83] | 57 [25; 82] | 8 [4; 17] | <0.0001 |
| Unknown | | 9 | 1 | 6 | 0 | |

[1] N (%) or Median (Q1, Q3)

[2] Pearson's Chi-squared test; Kruskal-Wallis rank sum test; Fisher's exact test

[a] Less than 40 copies of HIV per milliliter of blood

Supplementary table 2: Percent sensitivity and sensitivity of community-detected symptomatic and asymptomatic TB cases.

| | Community detected symptomatic TB | | Community detected asymptomatic TB | |
|---|---|---|---|---|
| | Sensitivity | Specificity | Sensitivity | Specificity |
| Primary definition | 56.7 | 69.4 | 51.1 | 69.4 |
| Alternative definition 1 | 60.0 | 69.4 | 57.6 | 69.4 |
| Alternative definition 2 | 66.7 | 69.4 | 58.8 | 69.4 |

#### SUPPLEMENTARY FIGURES

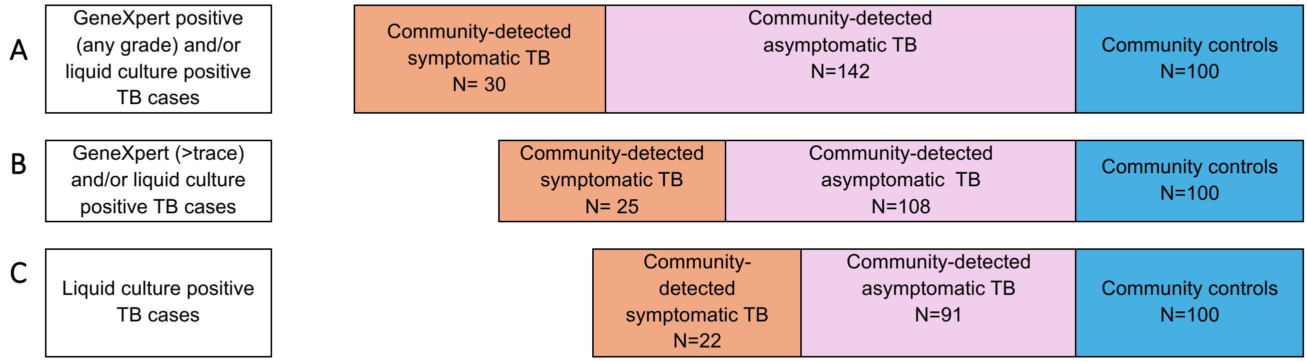

Supplementary Figure 1: Sensitivity analyses tested robustness of the primary analysis by using primary and alternative definitions of community-detected microbiologically-confirmed TB: (A) The primary definition of bacteriologically-confirmed TB includes all participants with sputum GeneXpert Ultra positive (any grade) and/or MGIT culture positive for Mtb; (B) Alternative definition 1 excludes people with GeneXpert Ultra “trace”-positive and culture-negative sputum and, (C) Alternative definition 2 includes only people with positive sputum by MGIT culture (regardless of GeneXpert Ultra status).

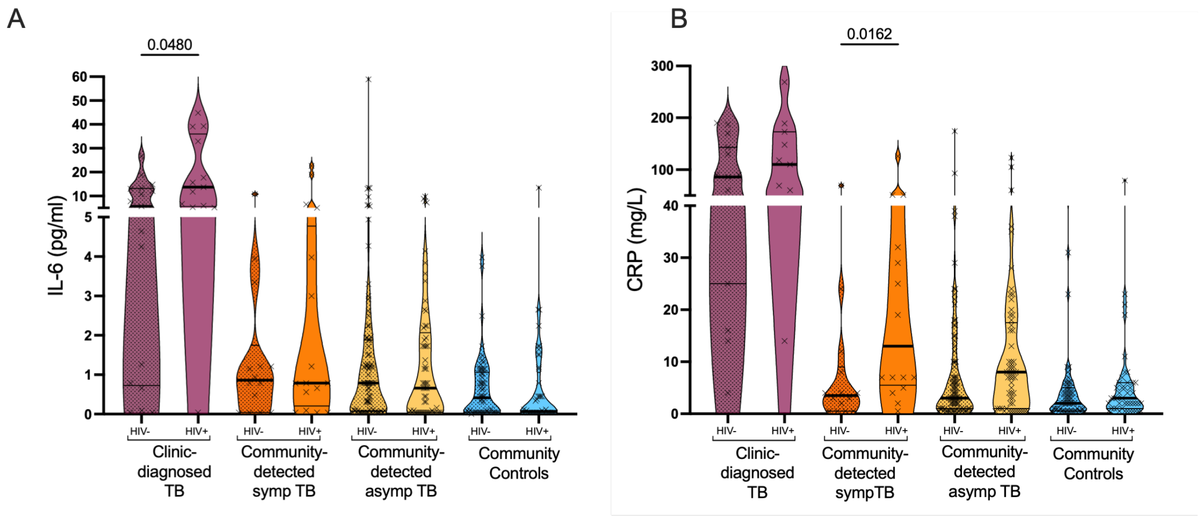

Supplementary Figure 2: Differential inflammatory plasma concentrations of IL-6 (A) and CRP (B) among people with clinic-diagnosed (maroon), community-detected symptomatic (ornage), community-detected asymptomatic (yellow) TB and community controls (blue) measured using luminex. Data is stratified by people without status HIV (dotted) and people living with HIV (clear). Data represents median values and IQR. P-values was obtained using Mann-Whitney U test for nonparametric dataset.

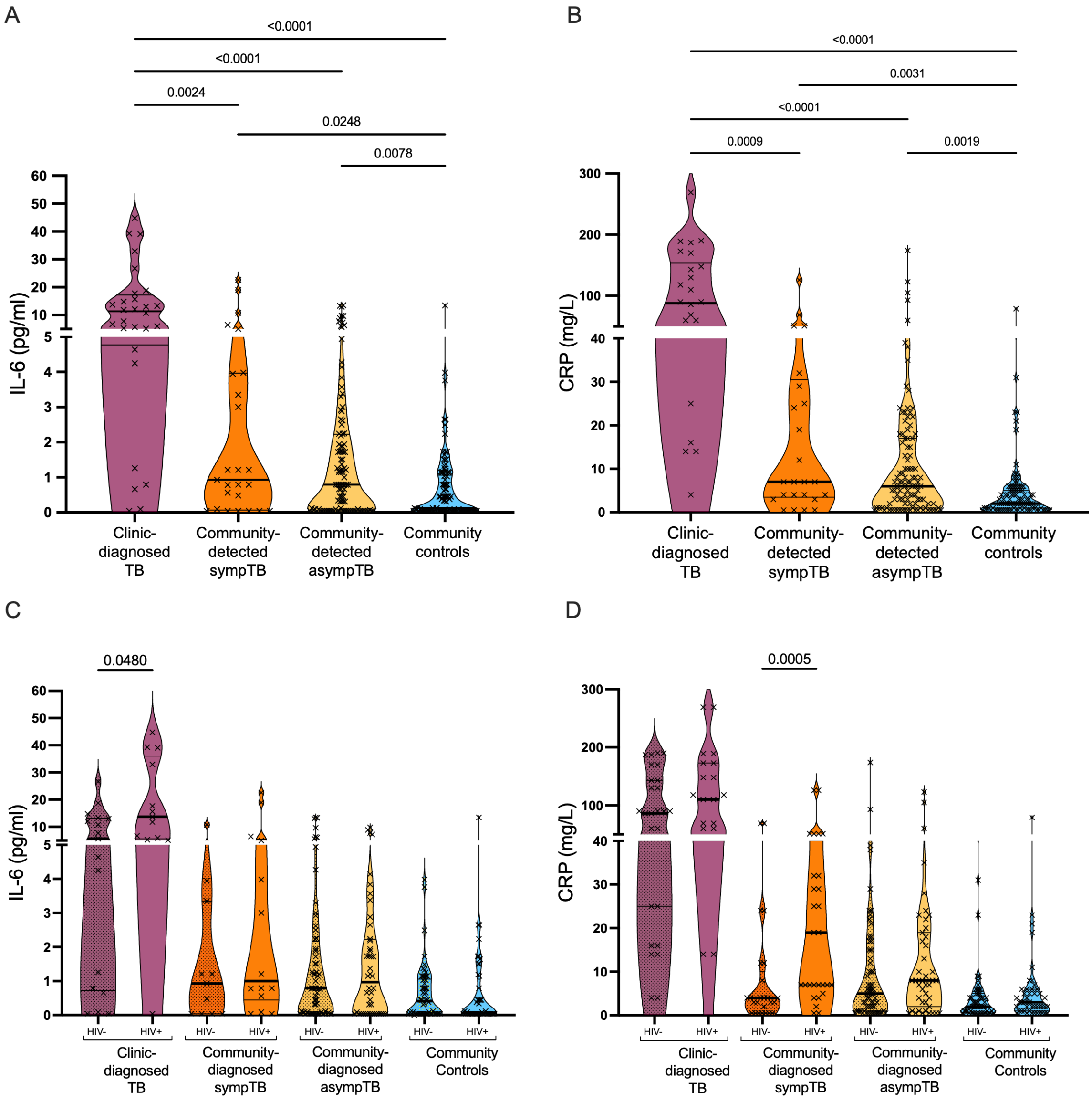

Supplementary Figure 3: Differential inflammatory plasma protein concentrations using alternative definition 1 (excluding people with GeneXpert Ultra “trace”-positive and culture negative sputum) for clinic-diagnosed TB (maroon), community-detected symptomatic TB (ornage), community-detected asymptomatic TB (yellow) and community controls (blue) . (A) IL-6 and (B) CRP concentrations were measured from plasma using luminex and stratified by people without status HIV (dotted) and people living with HIV (clear)(C-D). Data represents median values and IQR. P-values was obtained using Mann-Whitney U test for nonparametric a dataset.

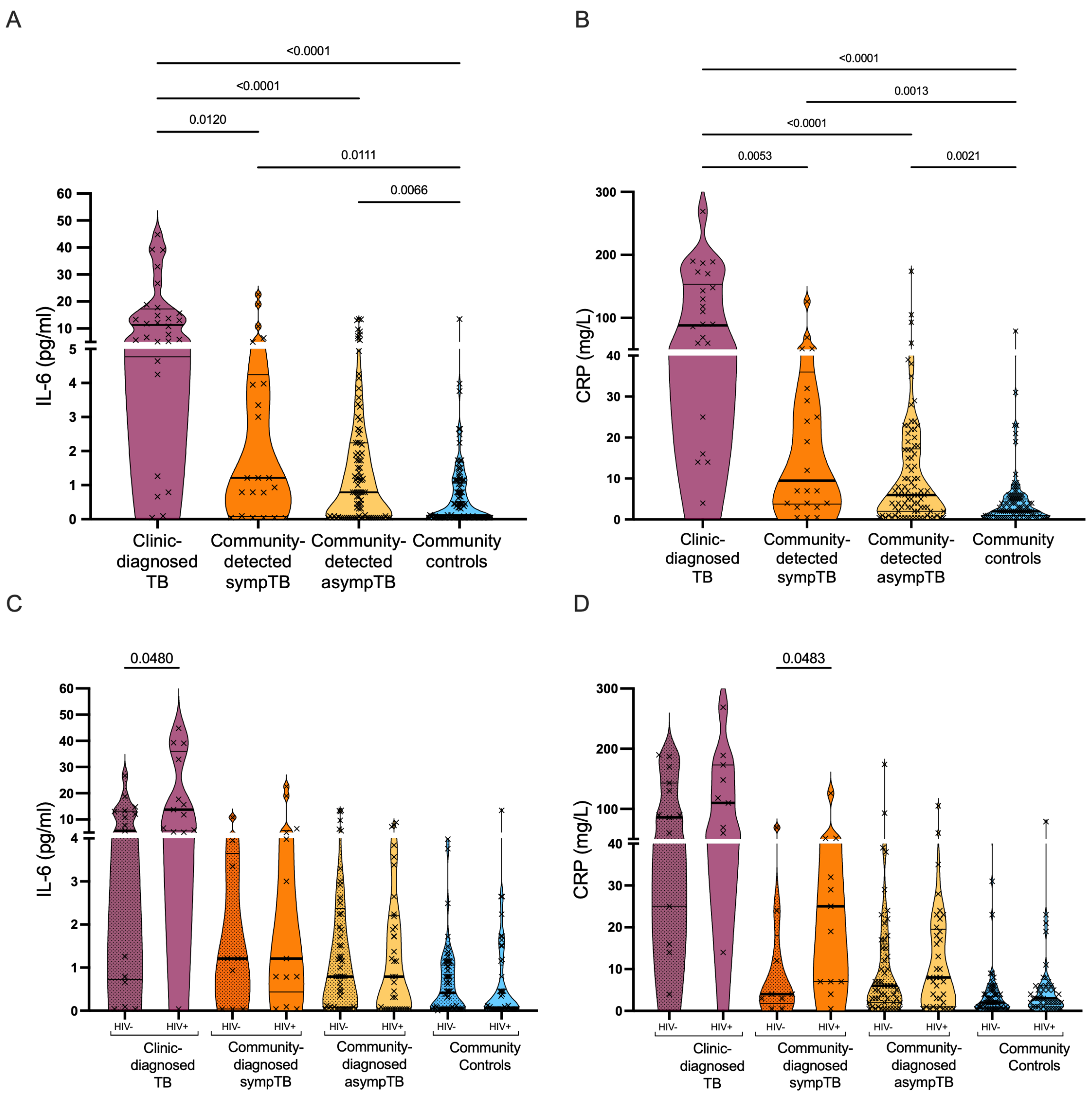

Supplementary Figure 4: Differential inflammatory plasma cytokine concentrations of (A) IL-6 and (B) CRP using alternative definition 2 (including only people with positive MGIT culture) for clinic-diagnosed TB (maroon), community-detected symptomatic TB (ornage), community-detected asymptomatic TB (yellow) and community controls (blue). Protein levels were measured from plasma using luminex and further stratified by people without status HIV (dotted) and people living with HIV (clear)(C-D). Data represents median values and IQR. P-values was obtained using Mann-Whitney U test for a nonparametric dataset.

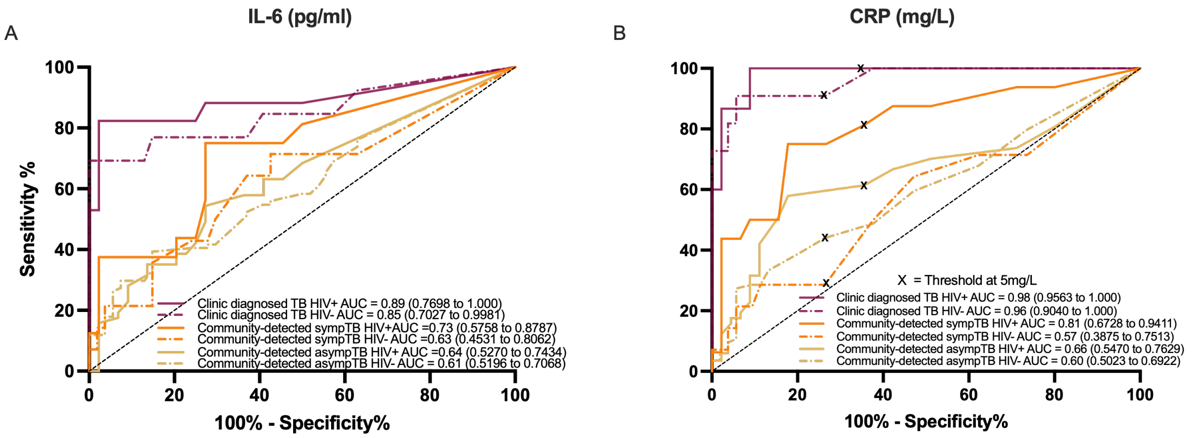

Supplementary Figure 5: Receiver operating characteristic curve (ROC) assessing performance of IL-6 (A) and CRP (B) in distinguishing clinic-diagnosed TB (maroon), community detected symptomatic (orange), subclinical TB cases (yellow) from community controls among people living with HIV (solid line) and people without HIV (dotted line). Data show the area under the curve (AUC) and the 95% confidence interval. 'X' indicates the sensitivity and specificity at a threshold of 5mg/L chosen based on the WHO's minimum requirement for a triage test.

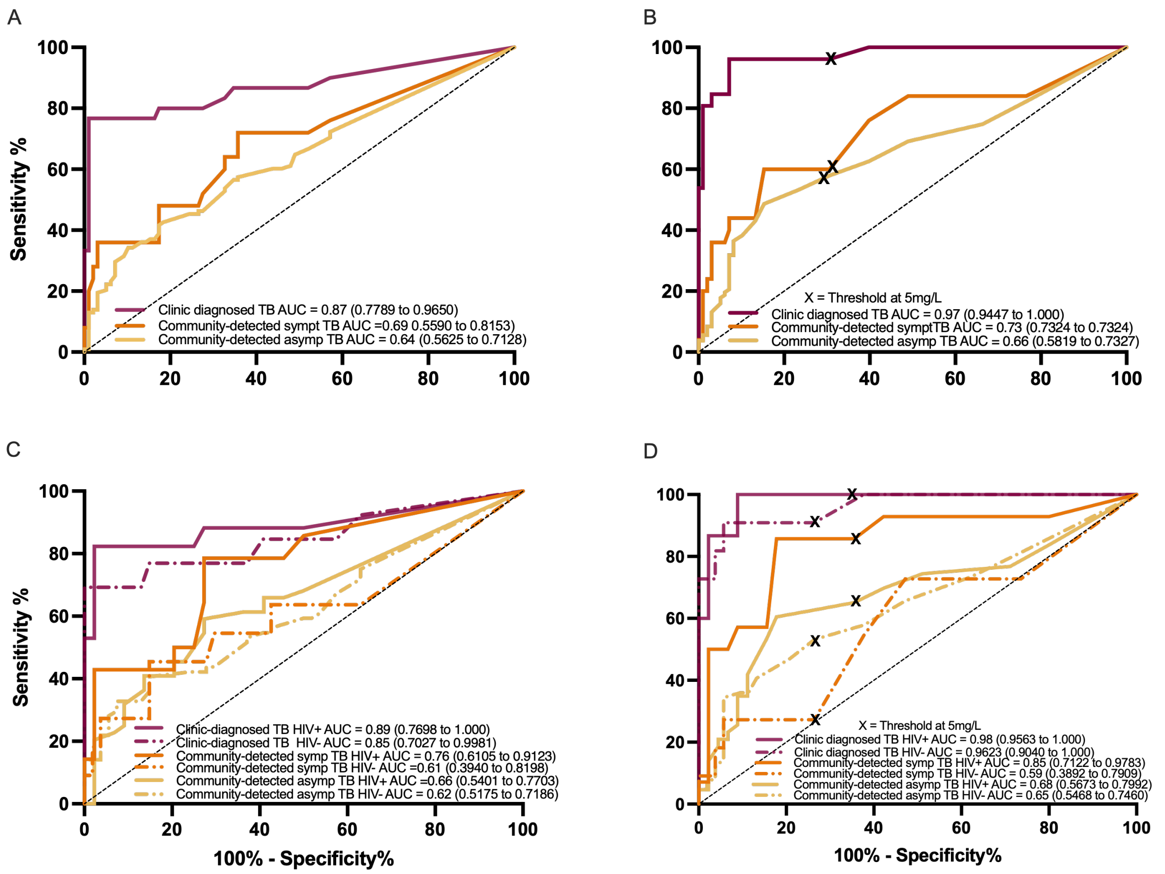

Supplementary Figure 6: Receiver operating characteristic curve (ROC) assessing performance of IL-6 and CRP using alternative definition 1 (excluding people with GeneXpert Ultra “trace”-positive and culture negative sputum) in distinguishing clinic-diagnosed TB (maroon), community detected symptomatic (orange), subclinical TB cases (yellow) from community controls. Data show the area under the curve (AUC) and the 95% confidence interval. ‘X’ represents cutoff of 5mg/L chosen based on the WHO’s minimum requirement for a triage test.

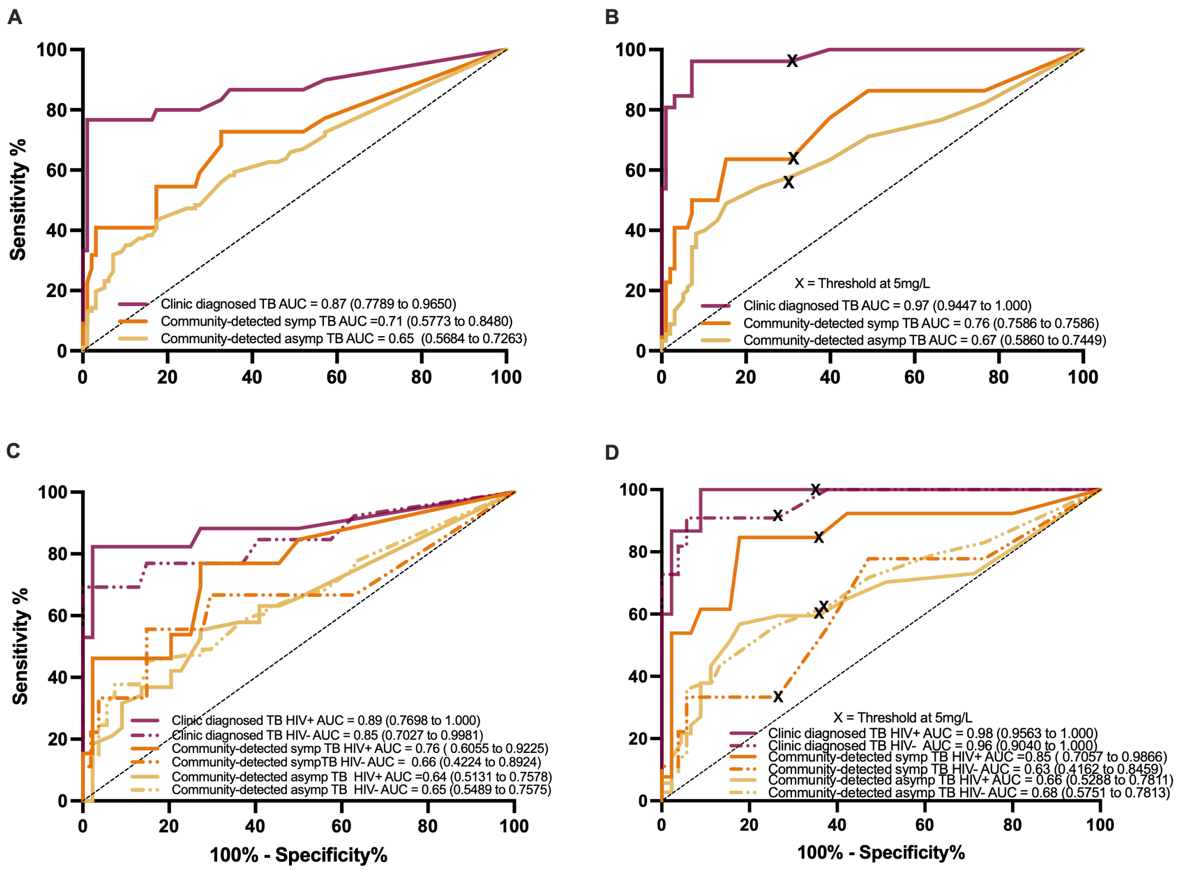

Supplementary Figure 7: Receiver operating characteristic curve (ROC) assessing performance of IL-6 and CRP using alternative definition 2 (including only people with positive MGIT culture) in distinguishing clinic-diagnosed TB (maroon), community detected symptomatic (orange), subclinical TB cases (yellow) from community controls. Data show the area under the curve (AUC) and the 95% confidence interval. ‘X’ represents threshold of 5mg/L chosen based on the WHO’s minimum requirement for a triage test.

A: Bacterial burden vs inflammatory markers

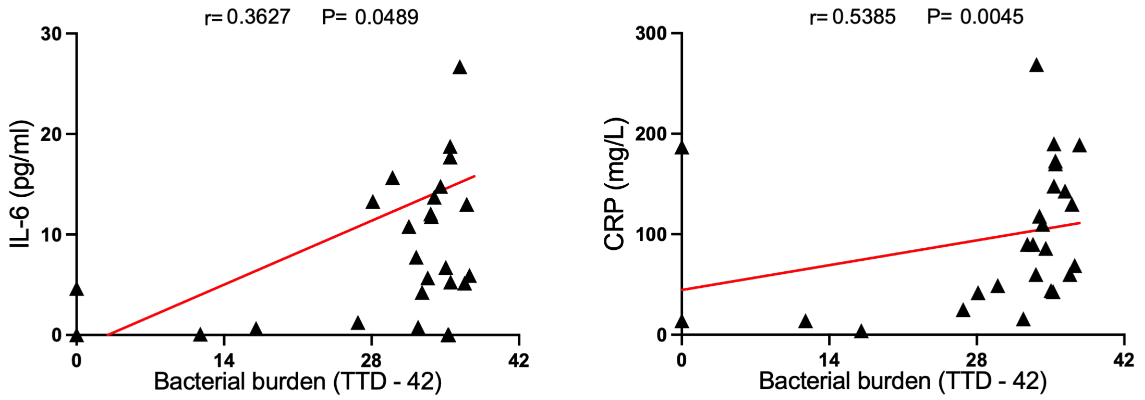

B: Extent of lung pathology vs inflammatory markers

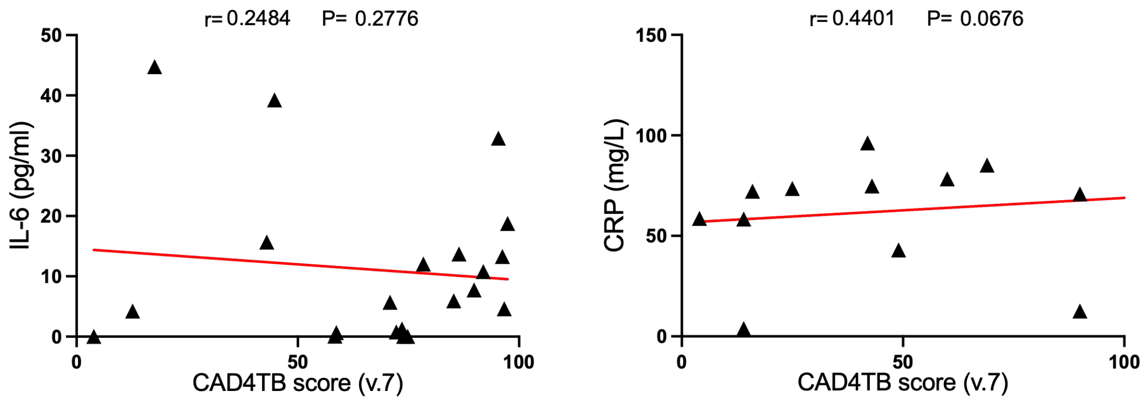

Supplementary Figure 8: Correlation analysis of IL-6 and CRP concentrations in clinic-diagnosed participants with (A) bacterial burden as measured by Time To Detection (days) minus 42 days and (B) extent of lung involvement, CAD4TBv7 scores, determined from digital chest radiography.

A: Bacterial burden vs inflammatory markers

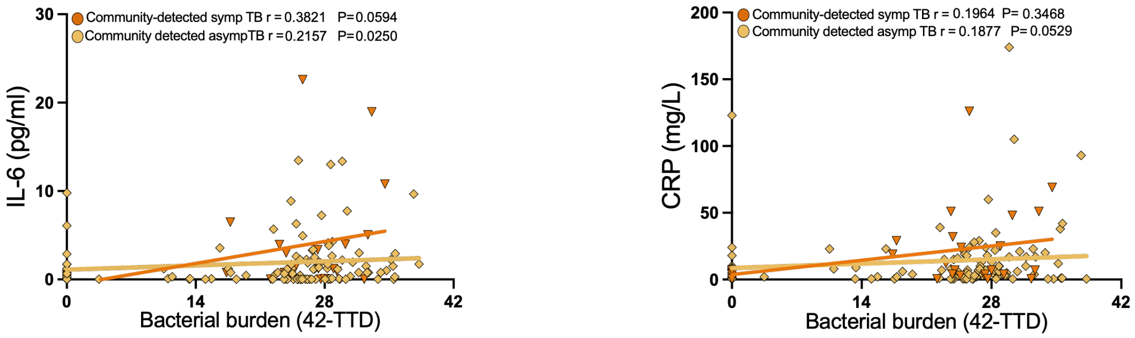

B: Extent of lung pathology vs inflammatory markers

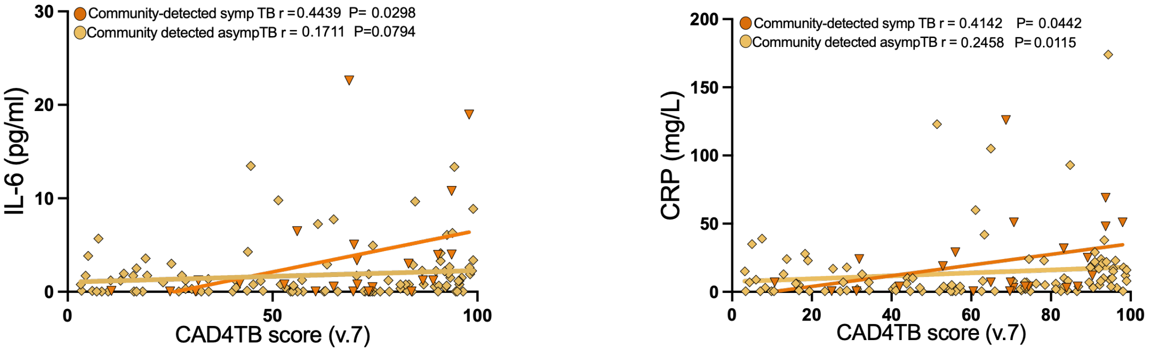

Supplementary Figure 9: Correlation analysis of IL-6 and CRP concentrations in community-detected symptomatic (blue) and subclinical (orange) TB participants with (A) bacterial burden as measured by Time To Detection (days) minus 42 days and (B) extent of lung involvement, CAD4TB scores, determined from digital chest radiography, using alternative definition 1 (excluding people with GeneXpert Ultra “trace”-positive and culture negative sputum) for community-detected TB groups).

A: Bacterial burden vs inflammatory markers

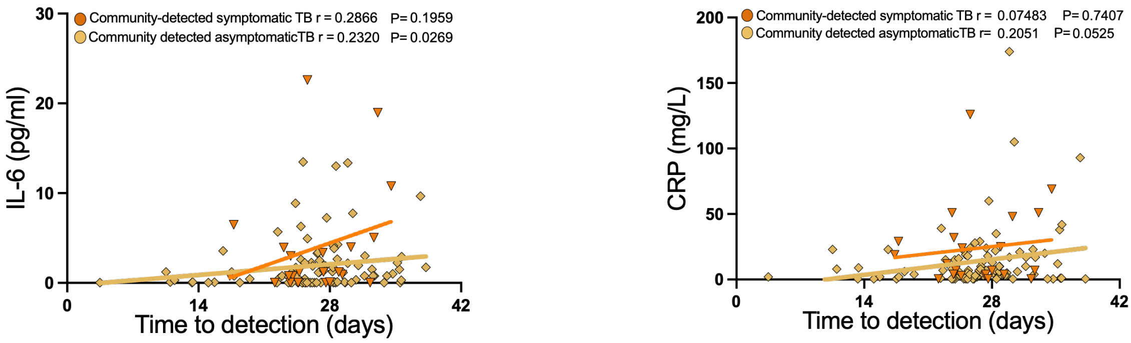

B: Extent of lung pathology vs inflammatory markers

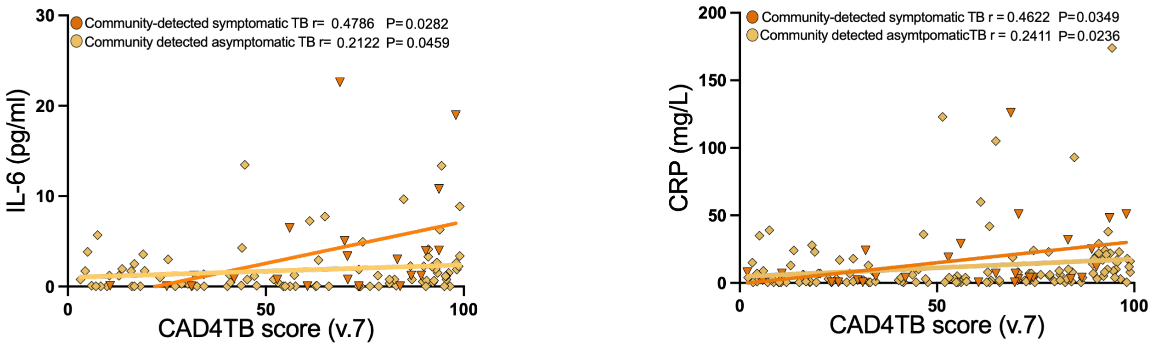

Supplementary Figure 10: Correlation analysis of IL-6 and CRP concentrations in community-detected symptomatic (blue) and subclinical (orange) TB participants with (A) bacterial burden as measured by Time To Detection (days) minus 42 days and (B) extent of lung involvement, CAD4TB scores, determined from digital chest radiography. Analyses were performed using alternative definition 2 (including only people with positive MGIT culture) for community-detected TB groups.
